# TB and HIV Drive Distinct and Separate Tissue Resident Memory Cell Subset Depletion

**DOI:** 10.64898/2026.02.12.26345105

**Authors:** Marjorie Nakibuule, Gift Ahimbisibwe, Marvin Martin Ssejoba, Rose Mulwana, Claire Precious Bisoboka, Marvin Joven Turyasingura, Josephine Nabulime, Febronius Babirye, Musana Abdusalaamu Kizito, Hervé Lekuya, Akello Suzan Adakun, Daisy Nalumansi, Irene Andia Biraro, Stephen Cose

## Abstract

**Background:** Tuberculosis (TB) and HIV co-infection cause profound immune dysregulation. Understanding how these infections alter immune cell distribution across systemic and tissue compartments is critical for improving therapeutic and vaccine strategies.

**Methods:** Flow cytometry was used to quantify CD4⁺ and CD8⁺ T cells, B cells, and tissue-resident memory (TRM) T and B cells in peripheral blood mononuclear cells (PBMCs), lung tissue, bronchoalveolar lavage (BAL), spleen, and lung-draining hilar lymph nodes (HLN) from individuals with pulmonary TB (PTB), disseminated TB (Diss TB), HIV only, or both TB and HIV infections.

**Results:** CD4⁺ T cell frequencies were significantly reduced in multiple compartments of HIV infected subjects, irrespective of TB status, indicating systemic immune suppression. CD8⁺ T-cell frequencies were elevated in the blood of HIV-infected individuals, suggesting a compensatory response to CD4⁺ T-cell loss. B-cell frequencies were reduced in PBMCs and lung tissue of TB subjects, regardless of HIV status. Notably, CD4⁺ TRM T cells were specifically depleted in lung tissue of HIV/TB co-infected individuals, whereas TRM B cells were selectively depleted in TB subjects, independent of HIV infection.

**Conclusion:** TB and HIV drive distinct and compartment-specific TRM cell loss in infected tissues. HIV primarily targets CD4⁺ TRM T cells, while TB specifically depletes TRM B cells, highlighting separate mechanisms of tissue-resident immune disruption. These findings emphasize the importance of tissue-specific immune analyses and provide new insights for targeted vaccine and immunotherapy strategies.

## INTRODUCTION

Following the COVID-19 pandemic, tuberculosis (TB) has returned to the top of the list of deaths from a single infectious disease (WHO, 2025). The 2025 WHO Global TB Report showed that approximately 10.7 million people contracted TB globally in 2024 and 1.23 million people died from TB (Estaji, Kamali, & Keikha, 2025; WHO, 2025). Uganda has long been among the 30 countries with the highest TB burden (G. WHO, 2020). Here, TB remains a major public health issue, exacerbated by factors such as high HIV co-infection rates, poverty, poor healthcare infrastructure, and limited access to diagnostics and treatment (Kirenga et al., 2015).

To combat the global TB epidemic, the World Health Organization (WHO) has set ambitious goals in its End TB Strategy, aiming to reduce TB deaths by 90% and the incidence rate by 80% by 2035. This strategy includes scaling up TB diagnosis and treatment and enhancing Bacille Calmette-Guérin (BCG) vaccination to protect children from TB (Savul & Duthie, 2023). However, the BCG vaccine, the only licensed vaccine for TB (Favorov et al., 2012; Setiabudiawan et al., 2022), has limited efficacy, particularly in adolescents and adults (Monteiro-Maia & Pinho, 2014) (Mangtani et al., 2014) in endemic settings, and who remain key transmitters of the disease (Tang, Yam, & Chen, 2016). Developing new vaccines requires a deeper understanding of the immunopathology of TB, especially at the site of infection in humans (G. Ahimbisibwe et al., 2023; Davids et al., 2020).

Historically, studies on human tissues have primarily focused on tissue pathology in various diseases, with only a few reports isolating and analysing tissue-resident immune cells (Kumar et al., 2017; Sathaliyawala et al., 2013). These studies, often conducted on organ donors, have identified key immune cell subsets, such as macrophages (Grassin-Delyle et al., 2020) (Snyder et al., 2021), NK cells, innate lymphoid cells (ILCs) (Ardain et al., 2019), tissue-resident memory T cells (TRM-T) (Thome & Farber, 2015) (Snyder et al., 2021), and mucosal-associated invariant T (MAIT) cells (Hinks et al., 2016). However, most studies in TB have relied on mouse and macaque models, which do not fully capture the complexity of the human immune response to TB. Only a handful of studies have examined immune cell subsets at site of infection of TB patients, with most focusing on bronchoalveolar lavage (BAL) (Ekberg-Jansson, Arvå, Nilsson, Löfdahl, & Andersson, 1999) or pleural fluid (Nemeth et al., 2009) (Fu et al., 2016) and occasionally lymph nodes when requested by clinicians. While PET-CT scans have provided insights into disease progression at the infection site, they do not allow for the study of specific immune cell subsets (Vorster, Sathekge, & Bomanji, 2014).

Tissue-resident memory T cells (TRM-T) are a specialised subset of immune cells that permanently occupy non-lymphoid tissues such as mucosal surfaces, skin, and solid organs without recirculating through the blood or lymph (Li, Xiao, Li, & He, 2025) (Szabo, Miron, & Farber, 2019). This fixed positioning at common pathogen entry points enables them to deliver rapid, localised immune responses (Snyder et al., 2021). Unlike circulating central memory (TCM) and effector memory (TEM) T cells, TRMs remain anchored within epithelial or parenchymal niches (Plunkett et al., 2022), often in regions excluded from routine immune surveillance (Takamura, 2018).

In infectious diseases, TRM-T rapidly deploy effector functions producing IFN-γ, TNF, IL-2, releasing granzyme B and perforin, and recruiting other immune cells. They are critical for controlling respiratory viruses such as influenza (Pizzolla et al., 2018) (de Bree et al., 2007), mucosal infections such as herpes simplex virus (Iijima & Iwasaki, 2014)), HIV (Kiniry et al., 2018) and intracellular bacteria where both CD4⁺ (TRM-T4) and CD8⁺ (TRM-T8) TRM T cells enhance early bacterial control (Ogongo et al., 2019; Yang et al., 2020; Yu et al., 2021). TRM-T4 contribute via IFN-γ, TNF, and IL-2 production, promoting macrophage activation and granuloma formation, while TRM-T8 exert cytotoxic effects to limit bacterial replication (Yang et al., 2020; Takamura et al., 2016).

In TB immunity, both TRM-T4 and TRM-T8 are uniquely adapted to remain in the lungs, where they are believed to offer rapid and localised protection against reinfection (Ogongo et al., 2021). This tissue residency is mediated by the expression of markers such as CD69 and CD103, which help anchor them within the tissue microenvironment (Plunkett et al., 2022).

Tissue-resident memory B cells (TRM-B) represent an emerging but equally important aspect of localised immunity. Unlike their circulating counterparts, TRM-B are retained within tissue and are poised for immune response upon antigen recall (Tan et al., 2022) (Allie et al., 2019). In TB, B cell frequencies are often reduced in the blood but enriched in the lung, particularly within granuloma-associated lymphoid structures, suggesting active recruitment to the site of infection (Krause et al., 2024). These lung TRM-B populations exhibit unique transcriptional and phenotypic signatures compared to circulating memory B cells, reflecting their adaptation to the lung microenvironment (Tan et al., 2022). Functionally, TRM-B cells can rapidly differentiate into antibody-secreting cells and are strategically positioned at the lung mucosa to provide immediate humoral defence against *M. tuberculosis (Krause et al., 2024).* Together, these findings highlight the critical but underexplored contribution of both TRM-T and TRM-B populations to TB immunity.

While animal models, *ex vivo* donor lungs and lung resections from TB patients have offered important insights, these approaches do not fully capture the immune landscape of natural infection. In addition, no studies have comprehensively characterised TRM-T and TRM-B cells across different clinical forms of TB (such as pulmonary, extrapulmonary, and TB/HIV co-infection), nor compared their distribution across multiple tissue sites. Addressing this gap is essential to understanding how resident immune populations shape TB pathogenesis and may inform novel vaccine or therapeutic strategies.

In this context, we undertook a human postmortem study to analyse immune cell subsets, location, phenotype, and function in tissues from TB subjects compared to non-TB subjects (G. Ahimbisibwe et al., 2023). Here, we show that TB and TB/HIV co-infection independently regulate the TRM microenvironment in the lungs of infected patients.

## MATERIALS AND METHODS

### ETHICS

This study received ethical approval from five ethics committees: the Makerere University School of Biomedical Sciences Research and Ethics Committee (SBS-REC-721), the Mulago National Referral Hospital Ethics Committee (MHREC 1849), the Kiruddu National Referral Hospital School of Biochemical Research and Ethics Committee (CRD/ADMIN/120/1), the Uganda National Council for Science and Technology Ethics Committee (HS703ES), and the London School of Hygiene and Tropical Medicine Ethics Committee (22922).

### CONSENTING

The Next-of-Kin (NoK) of the deceased was identified in accordance with Ugandan law. A trained grief counsellor explained the study and postmortem procedures to the NoK, allowing the NoK to ask questions and emphasizing that participation was voluntary. Written consent was obtained for a full autopsy, sample storage, and genetic testing.

### STUDY PARTICIPANTS

We recruited deceased subjects from Mulago National Referral Hospital (MNRH), Kampala Uganda, between January 2021 and June 2022. Active TB subjects were recruited from Wards 4, 5 and 6 of MNRH. Non-TB control subjects were road traffic accident victims recruited from the Surgical Emergency Unit (SEU) of MNRH. The NoK was approached at the SEU if the subjects’ medical records before death indicated normal parameters for physical examination, radiological and laboratory investigations. We excluded subjects with chest involvement and any other identified or known underlying chronic illness such as cancer.

Basic demographic characteristics of all subjects are presented in Table 1.

**Table 1.** Demographic and clinical characteristics of postmortem study participants. Participant recruitment and characteristics from the Surgical Emergency Unit (SEU) and Tuberculosis (TB) wards at Mulago National Referral Hospital. ^a^Surgical emergency unit ^b^TB wards ^c^Represents percentage ^d^median of the age group

|  |  | HOSPITAL WARD |  |
| --- | --- | --- | --- |
|  |  | SEU<sup>a</sup> | TB Wards<sup>b</sup> |
|  |  | 31 (46)<sup>c</sup> | 36 (54)<sup>c</sup> |
| GENDER | Male | 24(77)<sup>c</sup> | 24 (70)<sup>c</sup> |
|  | Female | 7(23)<sup>c</sup> | 11(30)<sup>c</sup> |
| AGE RANGE |  |  |  |
|  | 18-35 | 15 (27)<sup>d</sup> | 13(29)<sup>d</sup> |
|  | 36-55 | 10 (40)<sup>d</sup> | 15 (45)<sup>d</sup> |
|  | ≥ 56 | 6 (69)<sup>d</sup> | 7 (69)<sup>d</sup> |
| HIV STATUS | Positive | 8 (22)<sup>c</sup> | 24 (74)<sup>c</sup> |
|  | Negative | 23 (78)<sup>c</sup> | 12(26)<sup>c</sup> |
| CAUSE OF DEATH |  |  |  |
| Trauma |  |  |  |
|  | Head Trauma | 20 |  |
|  | Abdominal | 6 |  |
|  | Gunshot | 1 |  |
| non trauma |  |  |  |
|  | Intestinal Obstruction | 3 |  |
|  | Duodenal Ulcers | 1 |  |
| TB |  |  |  |
|  | Pulmonary TB |  | 12 |
|  | Extra Pulmonary TB |  | 13 |
|  | HIV Associated |  | 11 |

### TISSUE AND SAMPLE COLLECTION

The study pathologist carried out a full body postmortem on all study subjects to determine the cause of death and any underlying medical conditions not indicated in the files or medical records. To obtain tissue during the postmortem process, ∼5 cm^3^ of tissue was excised from the body and transferred to a 50ml tube containing RPMI. Samples were stored on ice for transfer to the BSL3 TB lab at the MRC/UVRI and LSHTM Uganda Research Unit, and samples processed as previously described (Gift Ahimbisibwe et al., 2023; Ahimbisibwe et al., 2025)

### T-SPOT.TB ASSAY

This assay was carried out using the T-SPOT.*TB* kit from Oxford Immunotec (Oxford, UK) following the manufacturer’s instructions, with the exception of incubation time which was increased to 48 hours to obtain repeatable results (Gift Ahimbisibwe et al., 2023). TB negative subjects were considered negative (TB^-^) if negative for the T-SPOT.*TB* assay.

### CELL PHENOTYPIC ANALYSIS BY FLOW CYTOMETRY

A 29-colour antibody T and B cell panel was used to phenotype the cells isolated from the various tissues. The panel consisted of: CD103 (BUV395), CD25 (BUV496), CD196 (BUV563), CD278 (BUV661), IgM (BUV737), CD45RA (BUV805), CD5 (BV421), CD3 (PACIFIC BLUE), CD38 (BV510), CCR7 (BV605), PD-1 (BV650), CD69 (BV711), CD28 (BV750), CD27 (BV785), CD24 (FITC), CD8 (SPARK BLUE 550), CD183 (BB700), CD10 (PerCP efluor710), FCLR4 (PE), CD127 (SPARK YG581), IgD (PE-Dazzle594), CD4 (PECY5), CD57 (PECY7), IgG (APC), KLRG1 (ALEXAFLUOR 647), CD21 (ALEXAFLUOR 700), CD19 (APC-H7), HLADR (APC-FIRE 810) and LIVE/DEAD (ZOMBIE UV) (BioLegend).

### PHENOTYPIC STAINING

Viability staining was performed with the fixable viability dye, Zombie UV, for 20 minutes in the dark at room temperature. Cells were washed and resuspended in 100mL of surface antibody cocktail for 20 minutes in the dark at 4^°^C, after which they were washed to remove excess antibody. Cells were acquired using a Cytek Aurora 5-laser spectral flow cytometer. Controls to remove spectral overlap and FMOs were used to establish gates. All flow cytometry data was analysed using FlowJo version 10.8.1. The gating strategy to identify T and B cell subsets in PBMCs and tissue (LUNG) is shown in Figures 1 and 2, respectively.

**Figure 1.**
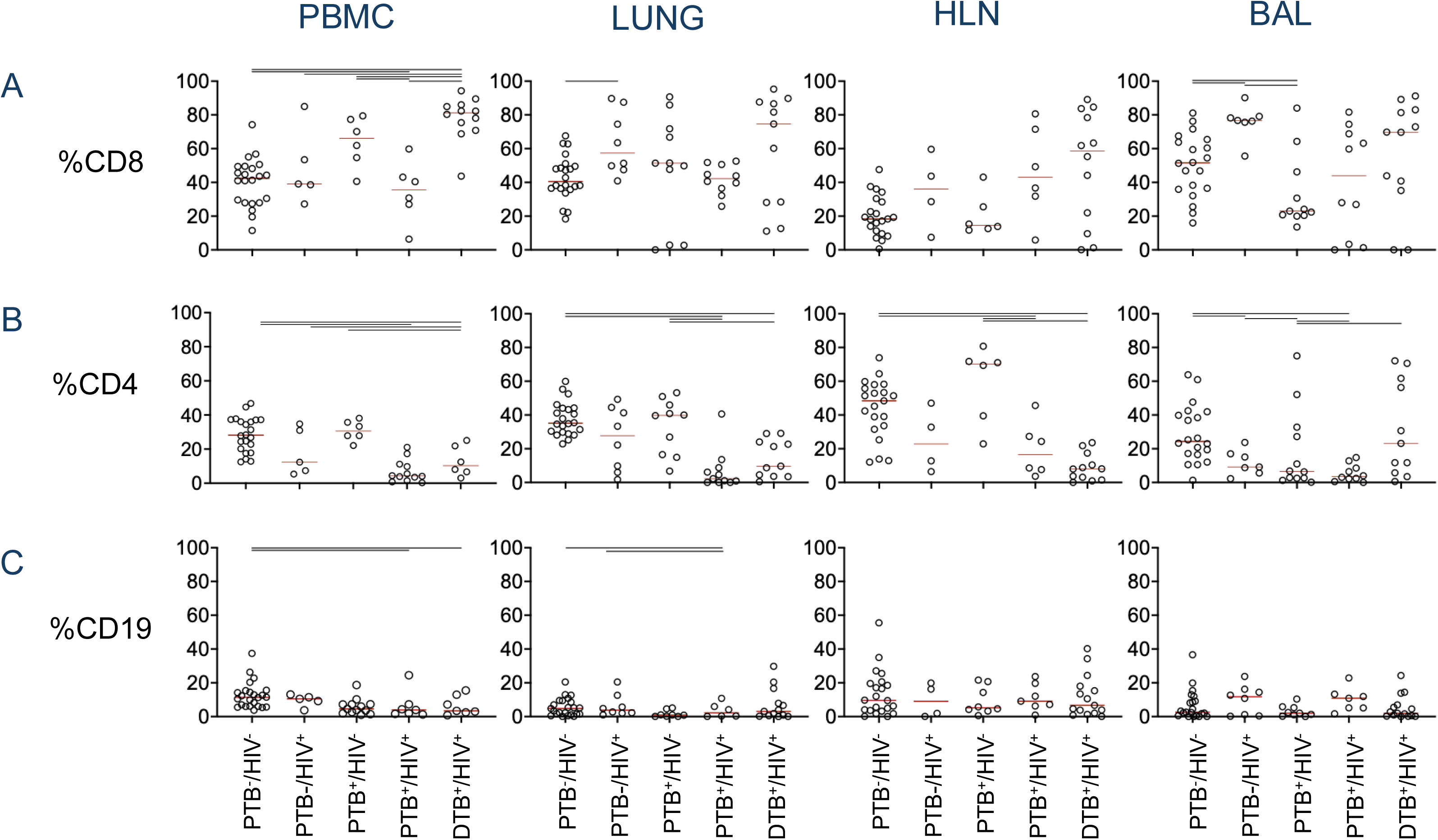
Gating strategy for T and B cell subsets in PBMCs. Representative flow cytometry plots illustrating the sequential gating strategy for identifying T and B cell populations in peripheral blood mononuclear cells (PBMC). Lymphocytes were first selected based on forward scatter (FSC-A) and side scatter (SSC-A), followed by singlet discrimination using FSC-H versus FSC-A and live cell gating with Zombie UV viability dye. T cells were identified as CD3⁺ and further subdivided into CD4⁺ and CD8⁺ populations. CD4⁺ and CD8⁺ T cells were subsequently gated for the tissue-resident markers CD69 and CD103. B cells were identified as CD19⁺ and gated on CD10⁻CD21⁺ to select mature B cells. Class switched and memory B cell subsets were distinguished based on IgM and IgD expression, with IgM⁻/IgD⁻ class switched B cells further analysed for CD69 and CD103 expression.

**Figure 2.**
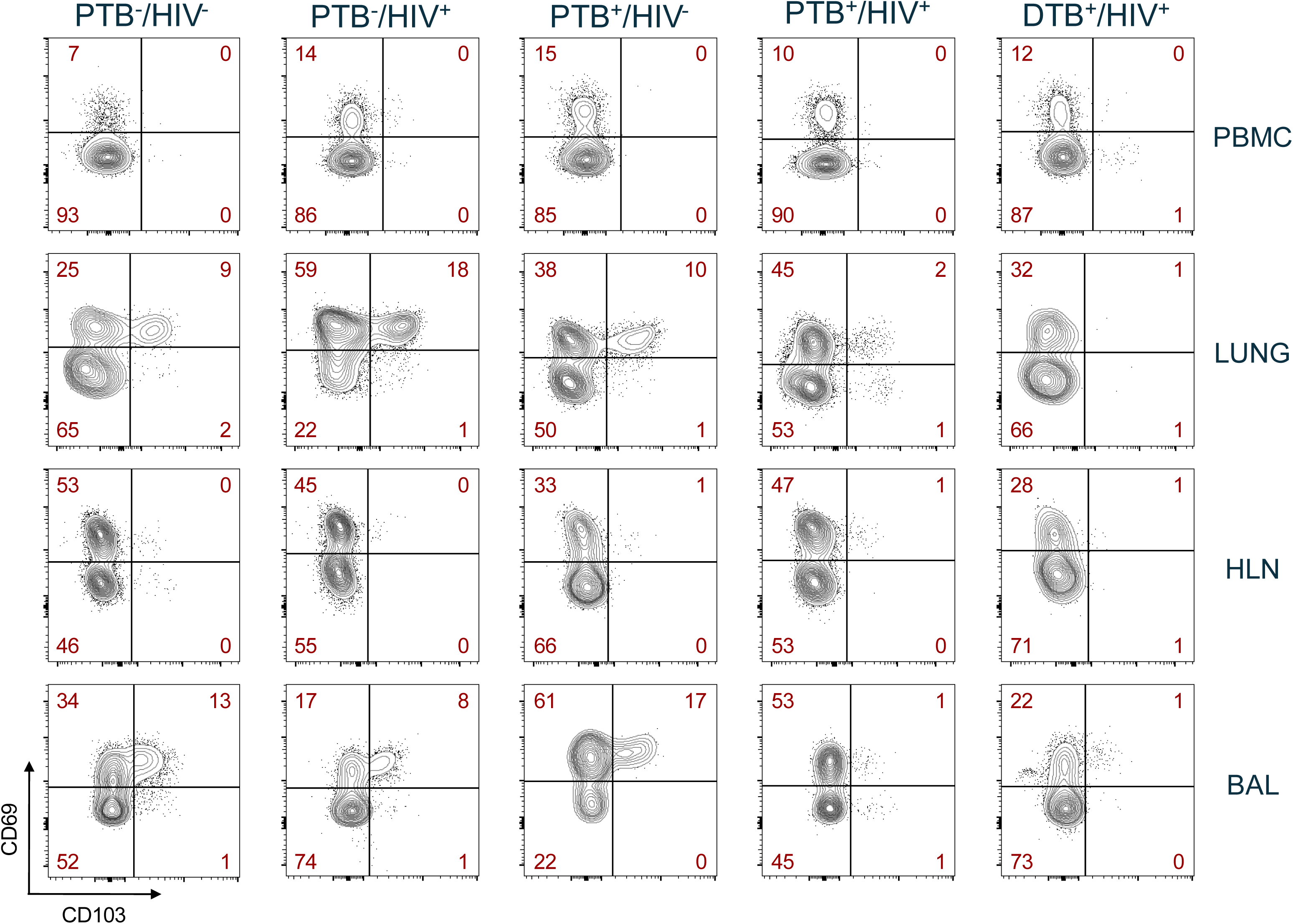
Gating strategy for T and B cell subsets in Lung tissue. Representative flow cytometry plots for lung-derived lymphocytes using a similar sequential gating strategy as in Figure 1. Lymphocytes were first selected based on forward scatter (FSC-A) and side scatter (SSC-A), followed by singlet discrimination using FSC-H versus FSC-A and live cell gating with Zombie UV viability dye. T cells were identified as CD3⁺ and further subdivided into CD4⁺ and CD8⁺ populations. CD4⁺ and CD8⁺ T cells were subsequently gated for the tissue-resident markers CD69 and CD103. B cells were identified as CD19⁺ and gated on CD10⁻CD21⁺ to select mature B cells. Class switched and memory B cell subsets were distinguished based on IgM and IgD expression, with IgM⁻/IgD⁻ B cells further analysed for CD69 and CD103 expression

### LUNG PERFUSION

Lung perfusion was performed in the mortuary. Briefly, a cadaver was positioned supine, and a chest incision was made to expose the thoracic cavity. The pulmonary vessels were carefully identified and cannulated *in situ*, followed by resection of the heart–lung block from the thoracic cavity. Perfusion was performed using a mixture of anti-CD45 FITC and Phosphate Buffered Saline (PBS) solution to achieve intravascular (i.v.) staining. Lung, blood perfusate, and bronchoalveolar lavage (BAL) samples were collected for downstream single-cell isolation, *ex vivo* staining, and flow cytometric analysis.

### STATISTICAL ANALYSIS

GraphPad prism version 8 software was used for graphical representation and statistical analysis.

## RESULTS

In this postmortem study, participants were recruited from both the Surgical Emergency Unit (SEU) and the Tuberculosis (TB) wards at Mulago National Referral Hospital (MNRH). Among the 31 individuals recruited from the SEU, 77% were male, with the majority aged between 18 and 35 years. Trauma-related injuries were the leading cause of death in this group (Table 1).

From the TB ward, 70% of the participants were male, with most aged between 36 and 55 years. Although the proportion of males in the TB ward was slightly lower compared to the SEU, it remained substantially higher than that of females. The primary cause of death in the TB ward was extra-pulmonary tuberculosis, often coexisting with HIV (Table 1).

### ANALYSIS OF CD4, CD8 AND CD19 FREQUENCIES ACROSS TISSUES

To asses general cell subset differences across tissues, we compared CD4^+^ and CD8^+^ T cell populations in uninfected control subjects (TSPOT-negative, HIV-negative; TB^-^/HIV^-^) with individuals who had HIV only (TB^-^/HIV^+^), pulmonary TB without HIV (PTB^+^/HIV^-^), pulmonary TB with HIV (PTB^+^/HIV^+^) and disseminated TB with HIV (DTB^+^/HIV^+^) (Figure 3).

**Figure 3.**
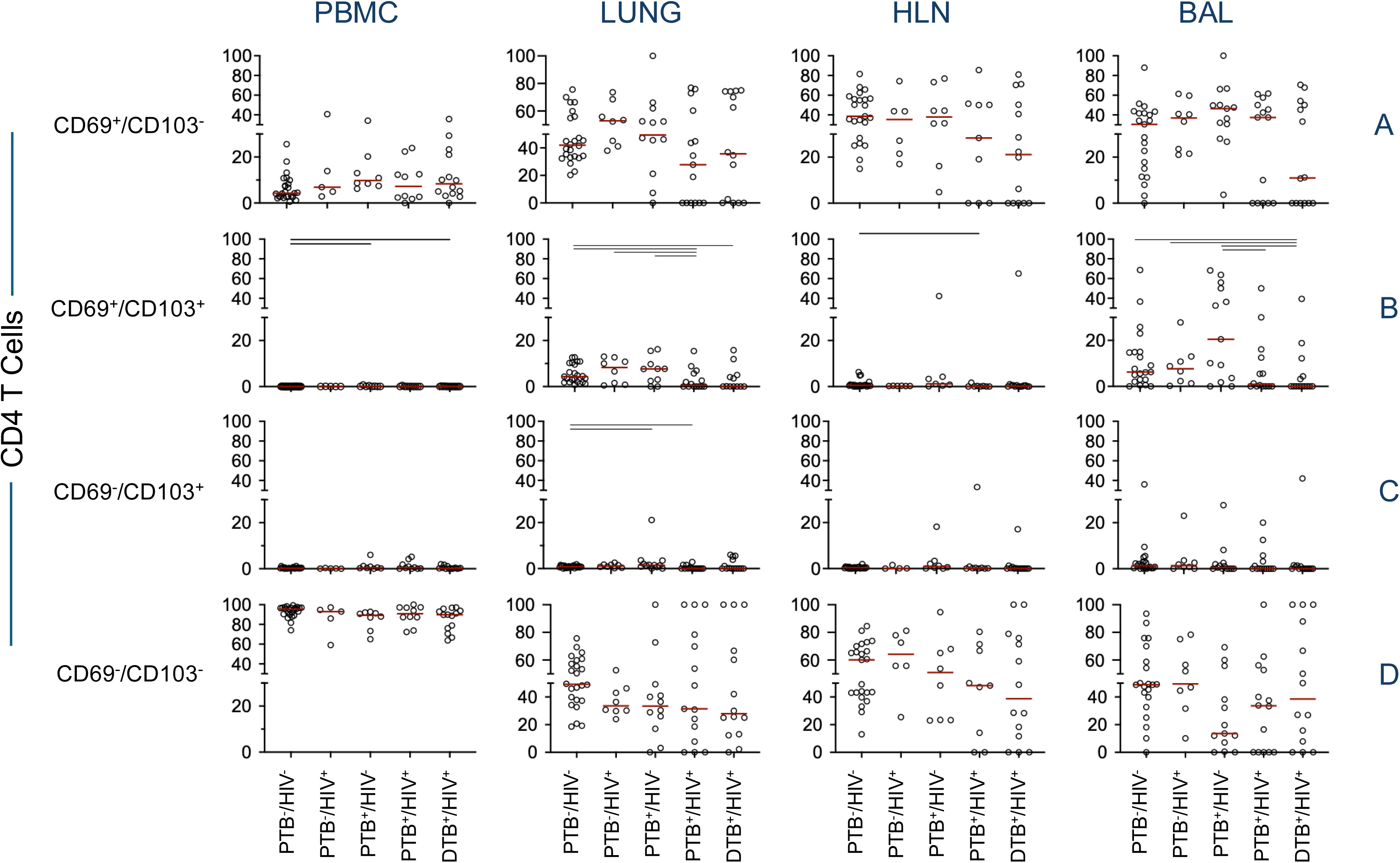
Distribution of lymphocyte subsets across tissues and patient groups. Percentages of (A) CD8⁺ T cells, (B) CD4⁺ T cells, and (C) CD19⁺ B cells in PBMC, LUNG, HLN and BAL samples from different patient groups. Red lines represent the median values for each group. Horizontal lines above the data indicate statistically significant differences between groups (p ≤ 0.05). PBMC, peripheral blood mononuclear cells; LUNG, lung tissue; HLN, lung draining hilar lymph node; BAL, bronchoalveolar lavage; PTB, pulmonary TB; DTB, disseminated TB.

When examining CD4 and CD8 T cell subsets across tissue or disease state, we found that CD4 T cells were depleted in all tissues in HIV-infected subjects, regardless of TB status (Figure 3A). Within PBMCs, PTB^-^/HIV^+^ subjects also had significantly lower CD4^+^ T-cell frequencies than DTB^+^/HIV^+^ subjects. In lung tissues, PTB^+^/HIV^+^ and DTB^+^/HIV^+^ subjects exhibited significantly lower CD4^+^ T-cell frequencies than TB^-^/HIV^+^ or PTB^+^/HIV^-^ subjects (Figure 3A). CD8 T cells were significantly elevated in the blood of subjects with disseminated TB (DTB^+^/HIV^+^; Figure 3B), with a similar trend across other tissues. CD8 T cells were significantly higher in the TB^-^/HIV^+^ subjects compared to the controls, perhaps indicating the loss of CD4 T-cells, but there were no significant differences across other HIV^+^ patient groups (Figure 3B).

When we looked at B cell frequencies, we saw a notable drop in B cells in PBMC and lung tissue from individuals with HIV and TB coinfection (PTB^+^/HIV^+^, DTB^+^/HIV^+^; Figure 3C), with a trend towards lower B cells in the PTB^+^/HIV^-^ subject group.

### ANALYSIS OF TISSUE RESIDENT MEMORY (TRM) CELLS IN TISSUES

Having established the differences in frequency of the surface markers CD4, CD8 and CD19, we next examined the TRM cell population in the various tissues. TRM cells were identified based on co-expression of CD69 and CD103, where CD69 serves as an activation and tissue ingress marker, and CD103 indicates tissue retention within the parenchymal tissues (Cibrián & Sánchez-Madrid, 2017).

For CD8^+^ T-cells, we saw an overall depletion of CD69^-^/CD103^-^ (non TRM) CD8 T cells in all subject groups compared to our uninfected controls (TB^-^/HIV^-^), with a consequent increase in CD69^+^/CD103^-^ T cells in PBMC, suggesting an increase in activated CD8^+^ T cells in the blood across all infection states (Supplementary Figures 1 and 2).

In contrast, HIV infection was associated with a specific reduction of CD69⁺/CD103⁺ CD4⁺ TRM T-cells in the lung and BAL of individuals with both TB and HIV, but not in those with either infection alone, indicating a profound and specific loss of TRM-T4 cells in lung tissue when both TB and HIV were present (Figures 4 and 5).

**Figure 4.**
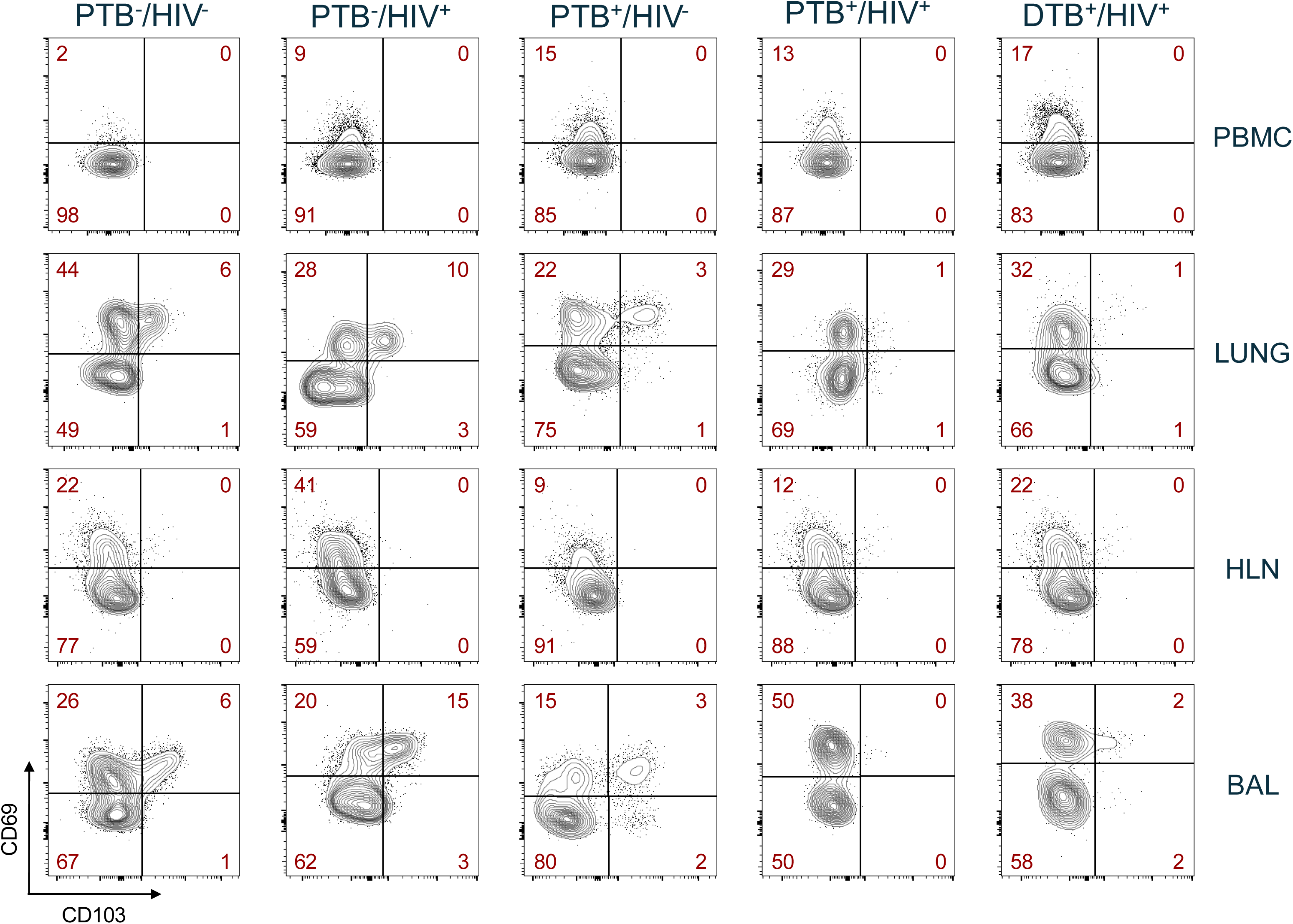
Representative staining of CD4^+^ TRM T cells across tissues and disease state. Representative flow cytometry contour plots showing CD69 and CD103 expression on CD4⁺ T cells from PBMC, LUNG, HLN and BAL. Rows indicate the tissue type, columns represent the different clinical groups. Red numbers represent the percentage of cells falling within each plot quadrant. PBMC, peripheral blood mononuclear cells; LUNG, lung tissue; HLN, lung draining hilar lymph node; BAL, bronchoalveolar lavage; PTB, pulmonary TB; DTB, disseminated TB.

**Figure 5.**
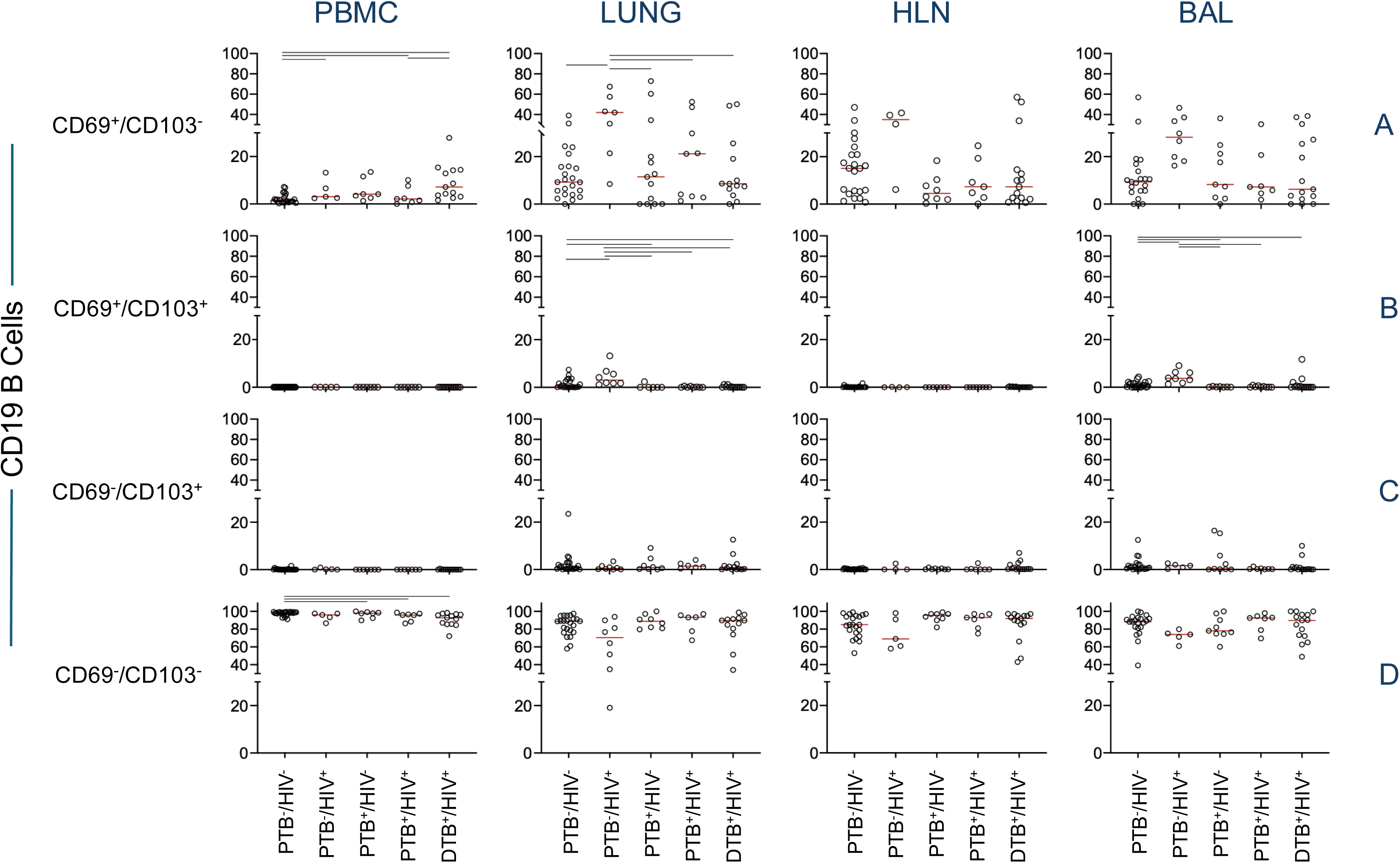
Frequency of CD69 and CD103 CD4⁺ T cell subsets across tissues and patient groups. Individual dots represent data from individual participants. Red horizontal lines indicate the median values for each group. Horizontal bars above groups denote statistically significant differences with p ≤ 0.05. Rows indicate the tissue type, columns represent the different clinical groups. PBMC, peripheral blood mononuclear cells; LUNG, lung tissue; HLN, lung draining hilar lymph node; BAL, bronchoalveolar lavage; PTB, pulmonary TB; DTB, disseminated TB.

In contrast to both CD4 and CD8 T cells, TB disease caused a depletion the B cell TRM subset (CD69^+^/CD103^+^) in the airways (Figure 6 and 7). This specific B cell subset depletion in TB subjects occurred regardless of HIV infection; in contrast, TRM-B cells remained significantly higher in TB^-^/HIV^-^ and TB^-^/HIV^+^ subjects, showing that this depletion is a TB-driven depletion event. Thus, TB/HIV coinfection, and TB infection itself drives specific and separate loss of TRM T and B cells in the airways of infected subjects.

**Figure 6.**
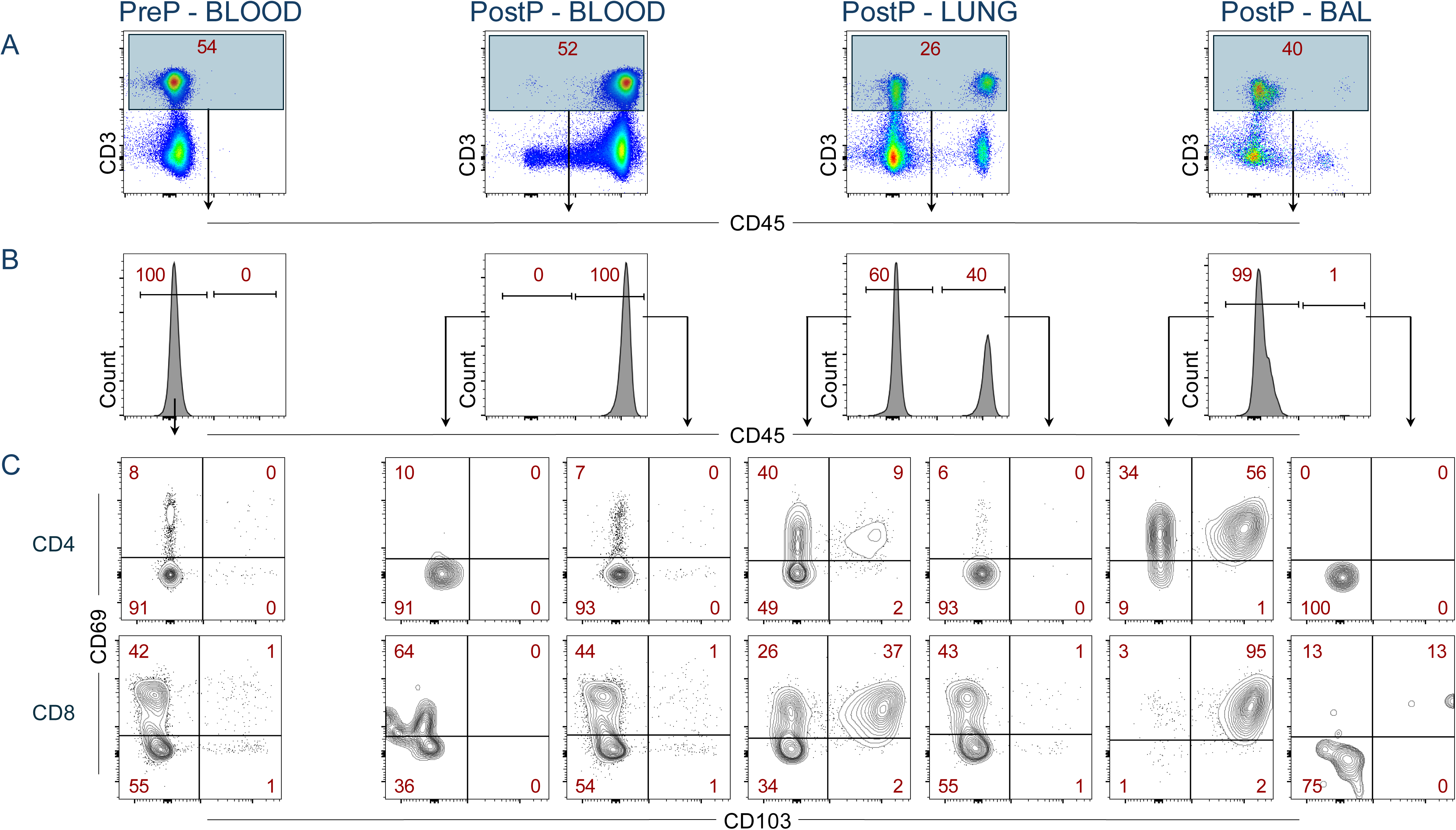
Representative staining of B cell TRMs across tissues and disease state. Representative flow cytometry contour plots showing CD69 and CD103 expression on B cells from PBMC, LUNG, HLN and BAL. Rows indicate the tissue type, columns represent the different clinical groups. Red numbers represent the percentage of cells falling within each plot quadrant. PBMC, peripheral blood mononuclear cells; LUNG, lung tissue; HLN, lung draining hilar lymph node; BAL, bronchoalveolar lavage; PTB, pulmonary TB; DTB, disseminated TB.

**Figure 7.** Frequency of CD69 and CD103 B cell subsets across tissues and patient groups. Individual dots represent data from individual participants. Red horizontal lines indicate the median values for each group. Horizontal bars above groups denote statistically significant differences with p ≤ 0.05. Rows indicate the tissue type, columns represent the different clinical groups. PBMC, peripheral blood mononuclear cells; LUNG, lung tissue; HLN, lung draining hilar lymph node; BAL, bronchoalveolar lavage; PTB, pulmonary TB; DTB, disseminated TB.

To verify that the lymphocyte subsets analysed were truly tissue resident, we assessed intravascular CD45-FITC staining after postmortem lung perfusion. Only a very small proportion of CD45-FITC–positive cells was detected within lung tissue, with staining largely confined to the vascular fraction. In contrast, the majority of TRM-T and TRM-B cells were CD45-FITC–negative (Figure 8), indicating that these populations were embedded within the lung parenchyma rather than circulating at the time of sampling. These findings confirm that the TRM subsets characterised in this study represent *bona fide* tissue-resident memory cells.

**Figure 8.** Identification of tissue-resident T cells in human lung tissue. Intravascular (i.v.) staining was performed using a postmortem human lung perfusion protocol. A control blood sample (pre-perfusion, PreP) was collected from the pulmonary vessels before perfusion with anti-CD45–FITC, and a second blood sample (post-perfusion, PostP) was collected after antibody perfusion. Lung (LUNG) tissue and bronchoalveolar lavage (BAL) samples were harvested post-perfusion, and single-cell suspensions were prepared for flow cytometric analysis. **(A)** CD3+ T cells were gated out from the selected tissue. **(B)** CD45 staining of CD3-gated cells (shadowed region in (A)) showing stain negative (CD45^-^) and positive (CD45^+^) T cell subsets. **(C)** CD4^+^ (upper panels) and CD8^+^ (lower panels) T cell populations were gated on CD45^+/-^ populations (B) and analysed for expression of CD69 and CD103. Percentage of gated cells in each region/quadrant are shown in red. PreP, pre-perfusion; PostP, post-perfusion; BAL, bronchoalveolar lavage.

## Discussion

Our data showed that CD4 T cell frequencies were significantly reduced across multiple compartments in PTB^+^/HIV^+^ and DTB^+^/HIV^+^ subjects compared to healthy controls (Figure 3). In PBMC, both PTB^+^/HIV^+^ and DTB^+^/HIV^+^ subjects exhibited lower CD4 T cell frequencies, with TB^-^/HIV^+^ individuals also showing depletion, highlighting the impact of HIV on circulating CD4 T cells (Okoye & Picker, 2013) (Yao et al., 2014). A similar pattern was observed in both lung tissue and BAL, where HIV/TB coinfected subjects (PTB^+^/HIV^+^ and DTB^+^/HIV^+^) had significantly lower CD4 T cell frequencies than other subject groups. Beyond the lungs, CD4 T cell depletion was also evident in lymphoid organs such as the HLN and spleen, indicating systemic immune suppression (Okoye & Picker, 2013) (Vidya Vijayan, Karthigeyan, Tripathi, & Hanna, 2017). Since these sites are crucial for generating adaptive immune responses, their depletion may further weaken the body’s ability to control infection (Bromley et al., 2024; Marino, El-Kebir, & Kirschner, 2011; Scholz & Kashuba, 2021).

In contrast, CD8^+^ T cell frequencies in PBMC were significantly elevated in DTB^+^/HIV^+^ and PTB^+^/HIV^+^ subjects compared to both healthy controls and PTB^+^/HIV^-^ individuals, suggesting a compensatory effect of HIV on CD8^+^ T cells due to the loss of CD4 T cells. DTB^+^/HIV^+^ subjects also exhibited higher CD8^+^ T cell frequencies than TB^-^/HIV^+^ individuals, indicating that *M.tb* dissemination further drives CD8^+^ T cell expansion beyond HIV infection alone (Figure 3A). This aligns with evidence that CD8^+^ T cells play a role in controlling bacterial load (Catalfamo et al., 2011; Chetty et al., 2015), suggesting that the expansion of CD8^+^ T cells in coinfected individuals may compensate for CD4^+^ T cell depletion, but is not sufficient to control TB disease.

Our study further revealed that B cell frequencies were significantly reduced in PBMC of PTB^+^/HIV^+^ and DTB^+^/HIV^+^ subjects compared to TB^-^/HIV^-^ controls, with a trend towards lower frequencies in PTB^+^/HIV^-^ individuals, indicating that active TB, regardless of HIV status, disrupts circulating B cell populations (Maglione & Chan, 2009) (Krause et al., 2024). In lung tissue, only PTB^+^/HIV^+^ subjects exhibited lower overall B cell frequencies. However, there was a trend for lower B cell frequencies in the PTB^+^/HIV^-^ and DTB^+^/HIV^+^ groups, suggesting that there may be specific population within the overall B cell subset that may be specifically depleted.

The above data suggested that there was some perturbations in both CD4 T and B cell subsets, and that perhaps this perturbation was being driven by the loss of specific CD4 T or B cell subsets. We therefore went on to examined tissue-resident memory (TRM) CD4 T (TRM-4T) and B (TRM-B) cells. TRMs are specialised immune cells that reside at the tissues and do not circulate in blood (Szabo et al., 2019). These cells express CD69 that provides retention signal (Cibrián & Sánchez-Madrid, 2017) and CD103 that is required for epithelial adhesion and also enable long term tissue residency (Hardenberg, Braun, & Schön, 2018). In the current study we found that CD69⁺/CD103⁺ CD4 TRM cells were not significantly depleted in the lungs of subjects with either TB or HIV single infection (Figures 4 and 5). However, in TB/HIV coinfected individuals, we observed a specific depletion of the CD4 TRM T cell subset at the site of infection. Although other studies had shown perturbations in CD4 T cell subsets in the lungs ((Corleis et al., 2019)((Costiniuk & Jenabian, 2014; Costiniuk, Samarani, Wang, Vigano, & Ahmad, 2024) we have specifically shown that it is the TRM-T4 subset that is specifically depleted in TB/HIV coinfected subjects. This subset is not altered in uninfected or singly infected subjects, and suggests a synergistic effect of both HIV and TB infections in driving TRM-T4 to the site of TB infection (the lungs) and becoming targets for HIV infection and subsequent cell loss. Although we cannot attribute a causal pathway to this cell loss, we suggest that in endemic countries, an individual may become infected with TB first, driving TRM-T4 cells into the lungs, where they become targets for the later acquisition of HIV through sexual contact. Although we have not performed antigen-specific assays in this paper, we suggest that this depletion is not TB-specific, but an overall depletion of TRM-T4 cells driven by the inflammation caused by TB disease (likely as a result of HIV infection), and the subsequent infection and depletion of this cell subset by HIV.

Perhaps more surprising than the loss of the TRM-T4 subset in TB/HIV coinfected subjects, was the complete loss of the TRM-B subsets in individuals infected with TB, regardless of their HIV state. While B cells have traditionally been underappreciated in TB immunity, increasing evidence supports their role in local immune regulation, including antibody production, antigen presentation, cytokine secretion, and the organisation of tertiary lymphoid structures within infected tissues (Ashenafi & Brighenti, 2022; Maglione & Chan, 2009). TRM-B cells may therefore contribute to early containment of *M. tuberculosis* at the site of infection by supporting local immune architecture and coordinating interactions with T cells and macrophages.

Here we have shown that this subset is a TB-specific cell subset depletion, as it occurs in PTB^+^/HIV^-^ individuals, along with coinfected individuals, but not in HIV only or uninfected individuals. This data suggests a previously unknown role for TRM-B cells in TB infection and control at the site of infection. Again, it is difficult to ascribe causality, but we would suggest the loss of control of *M.tb* infection may be exacerbated by this loss of TRM-B cells. Exactly how *M.tb* may be causing this specific loss of B cells is currently under active investigation, and several mechanisms may explain this depletion. TB is capable of remodelling lung microenvironments and forming organised granulomas that alter local niches required for B-cell retention (Lyu et al., 2024) (Ashenafi & Brighenti, 2022). Disruption of chemokines such as CXCL13 or CXCL12 may impair retention signals, causing TRM B cells to exit or fail to be maintained (Hansen et al., 2005). Chronic antigenic stimulation from *M.tb* and sustained inflammatory cytokines (e.g., IFN-γ, TNF) may induce apoptosis of TRM B cells (Hansen et al., 2005). Moreover, although B cells are not canonical *M.tb* host cells, tissue destruction from infected macrophages, necrotic foci, or granuloma-associated proteases can damage B-cell–supporting niches (Stewart et al., 2023). Despite these losses, activated TRM B cells may migrate to draining lymph nodes (Menares et al., 2019), possibly explaining preserved B cell numbers in HLN.

Our lung perfusion experiments provided additional insights into TRM dynamics. Perfusion allows the distinction between intravascular and tissue-resident populations, confirming that the observed TRM cells were genuinely resident in the tissue and not circulating cells trapped in the lung vasculature (Anderson et al., 2014). These experiments showed that the TRM-T and -B cell populations reside firmly within the lung parenchyma, reinforcing that their depletion in TB/HIV or TB-only contexts reflects true tissue-level loss rather than redistribution into circulation. Furthermore, perfusion allows us to measure TRM cells directly in the lung tissue, where we hope to identify which cell subsets are sensitive to local tissue damage caused by TB. We also anticipate that the TRM cell loss will vary across lung regions, with areas containing granulomas showing greater depletion. This would suggest that the more the tissue is damaged, the more TRM cells will be lost.

### Conclusion

Our data indicate that CD4 TRM subset depletion in the lungs is predominantly driven by TB/HIV coinfection, whereas TRM B-cell depletion is TB-driven and independent of HIV. Lung perfusion experiments strengthen these conclusions by confirming true tissue residency of the depleted populations. Together, these findings demonstrate that tissue-resident immune populations have distinct vulnerabilities to TB and HIV. They highlight the critical importance of examining immune responses directly within diseased tissue to fully understand pathogen-specific impacts on local immunity.

### Limitations

Postmortem studies are naturally limited by the unique demographics of people seeking healthcare. Most individuals dying on the TB Ward were TB and HIV co-infected, meaning a paucity of HIV uninfected individuals.

### Recommendations

Future studies building on our perfusion approach should investigate the functional capacity of tissue-resident T and B cells, particularly their responses to M.tb-specific antigens, to better define their contributions to TB pathogenesis. In addition, understanding the precise mechanisms by which M.tb drives the loss of TRM B cells and why this subset is uniquely vulnerable.

## Data Availability

The data that support the findings of this study are available from the corresponding author upon reasonable request, subject to ethical and data protection restrictions.

## Acknowledgements

We thank the administrations of Mulago National Referral Hospital and Kiruddu National Referral Hospital for their support and cooperation in this study. We are deeply grateful to the bereaved families who consented to the enrollment of their deceased relatives.

## Funding

The authors declare that financial support was received for the research, authorship, and/or publication of this article. This work was supported by NIH Contract 75N93019C00070 and was conducted at the MRC/UVRI and LSHTM Uganda Research Unit. The Unit is jointly funded by the UK Medical Research Council (MRC), part of UK Research and Innovation (UKRI), and the UK Foreign, Commonwealth and Development Office (FCDO) under the MRC/FCDO Concordat agreement, and is also part of the EDCTP2 programme supported by the European Union

## Conflict of interest

The authors declare that the research was conducted in the absence of any commercial or financial relationships that could be construed as a potential conflict of interest.

---

**Supplementary Figure 1. Representative staining of CD8^+^ TRM T cells across tissues and disease state.**

Representative flow cytometry contour plots showing CD69 and CD103 expression on CD8⁺ T cells from PBMC, LUNG, HLN and BAL. Rows indicate the tissue type, columns represent the different clinical groups. Red numbers represent the percentage of cells falling within each plot quadrant. PBMC, peripheral blood mononuclear cells; LUNG, lung tissue; HLN, lung draining hilar lymph node; BAL, bronchoalveolar lavage; PTB, pulmonary TB; DTB, disseminated TB.

**Supplementary Figure 2. Frequency of CD69 and CD103 CD8⁺ T cell subsets across tissues and patient groups.**

Individual dots represent data from individual participants. Red horizontal lines indicate the median values for each group. Horizontal bars above groups denote statistically significant differences with p ≤ 0.05. Rows indicate the tissue type, columns represent the different clinical groups. PBMC, peripheral blood mononuclear cells; LUNG, lung tissue; HLN, lung draining hilar lymph node; BAL, bronchoalveolar lavage; PTB, pulmonary TB; DTB, disseminated TB.

## Notes

### Competing Interest Statement

The authors have declared no competing interest.

### Author Declarations

The Research and Ethics Committee of the Makerere University School of Biomedical Sciences, the Ethics Committee of Mulago National Referral Hospital, the Research and Ethics Committee of Kiruddu National Referral Hospital, the Uganda National Council for Science and Technology Ethics Committee, and the Ethics Committee of the London School of Hygiene and Tropical Medicine

