## supplementary figures for "TB and HIV Drive Distinct and Separate Tissue Resident Memory Cell Subset Depletion": supp.pdf

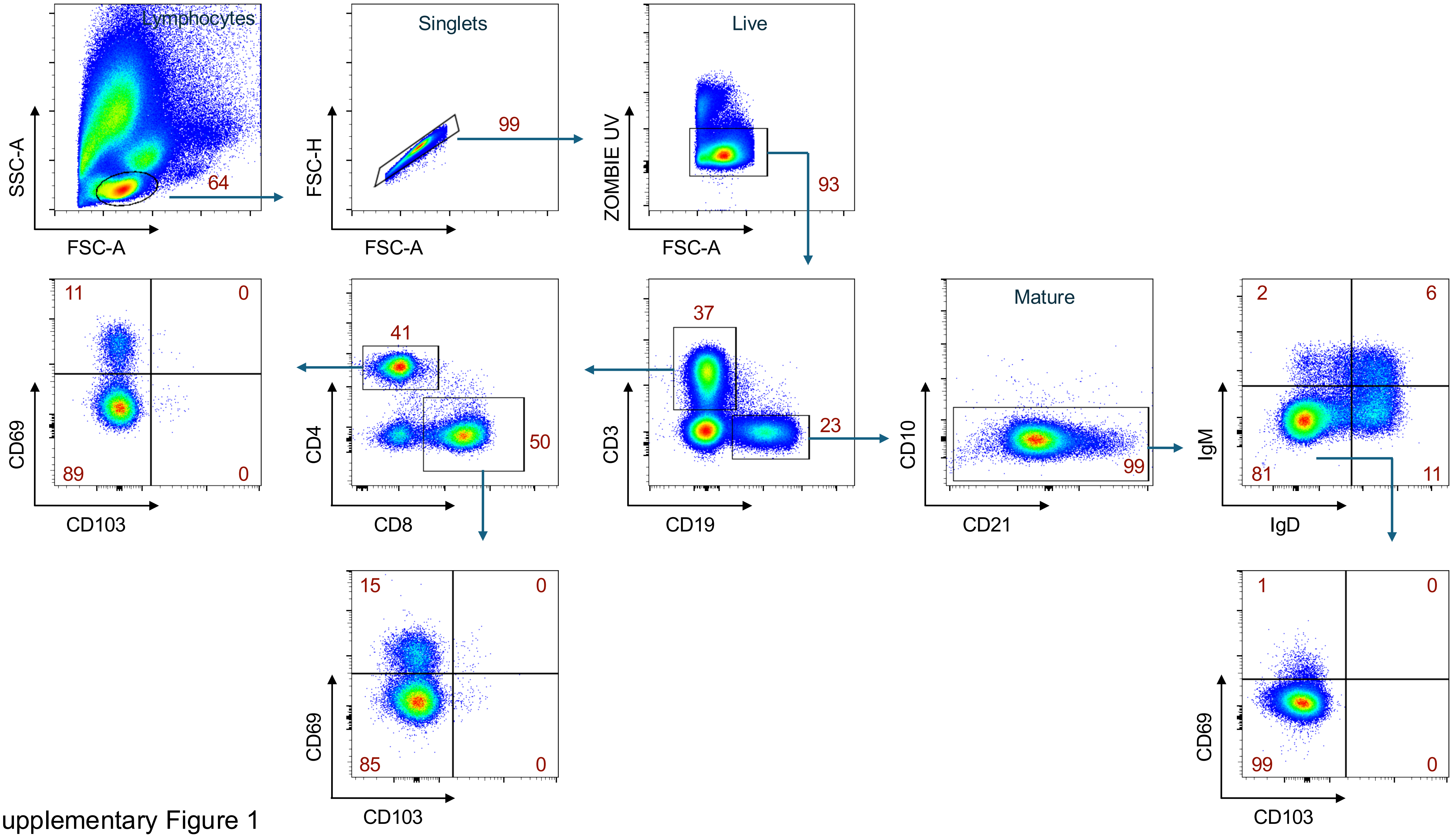


Supplementary Figure 1

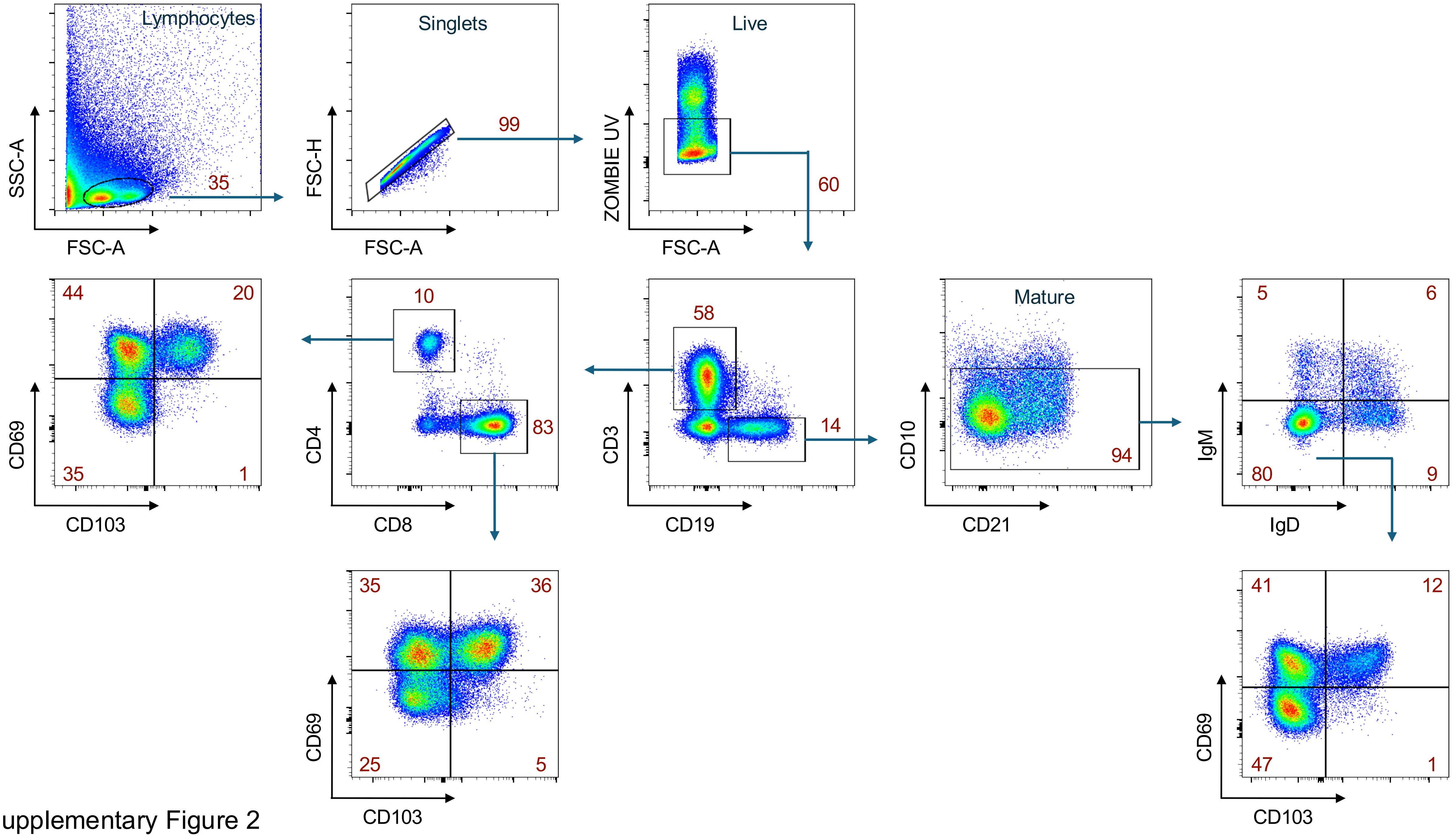


Supplementary Figure 2

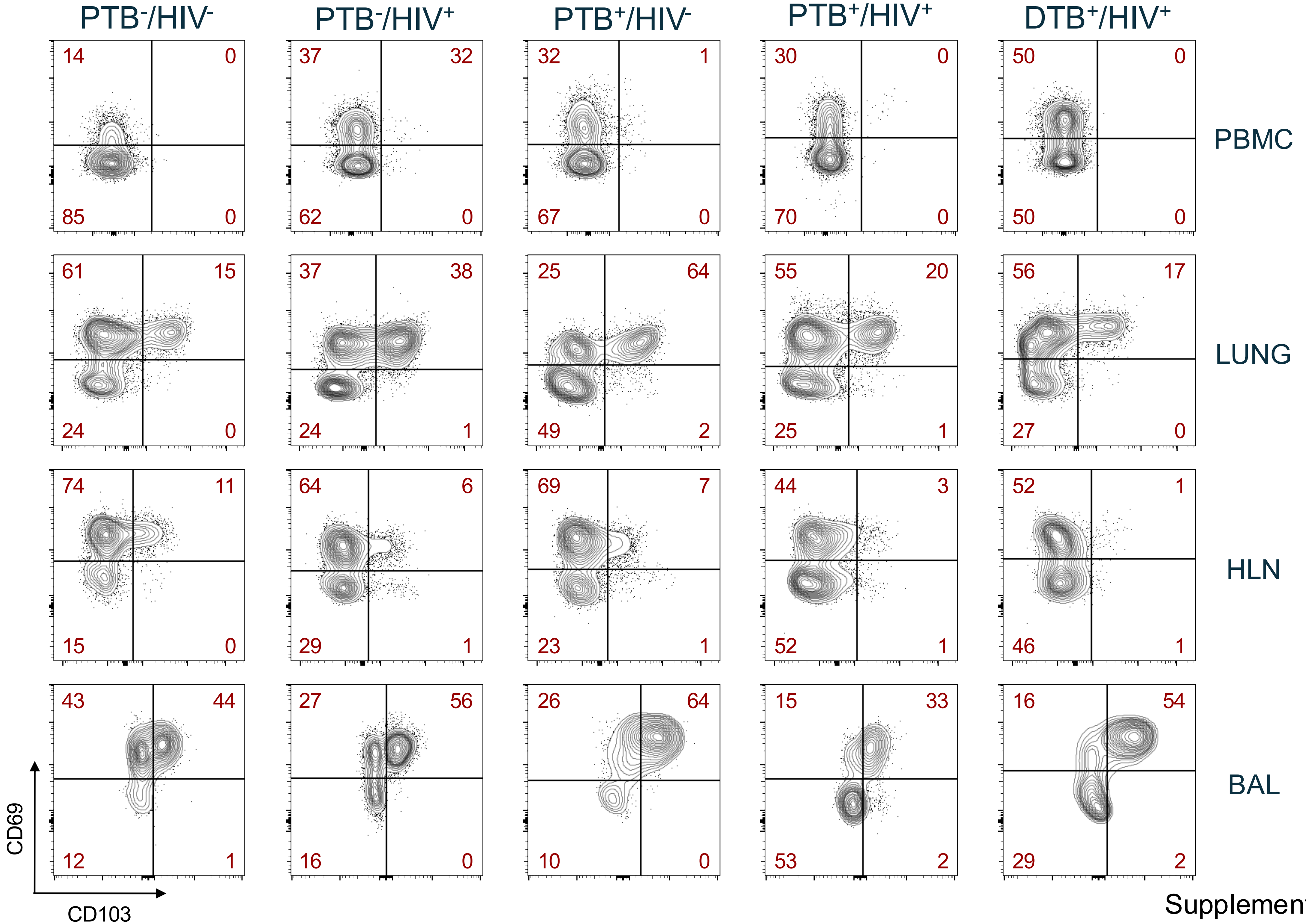


Supplementary Figure 3

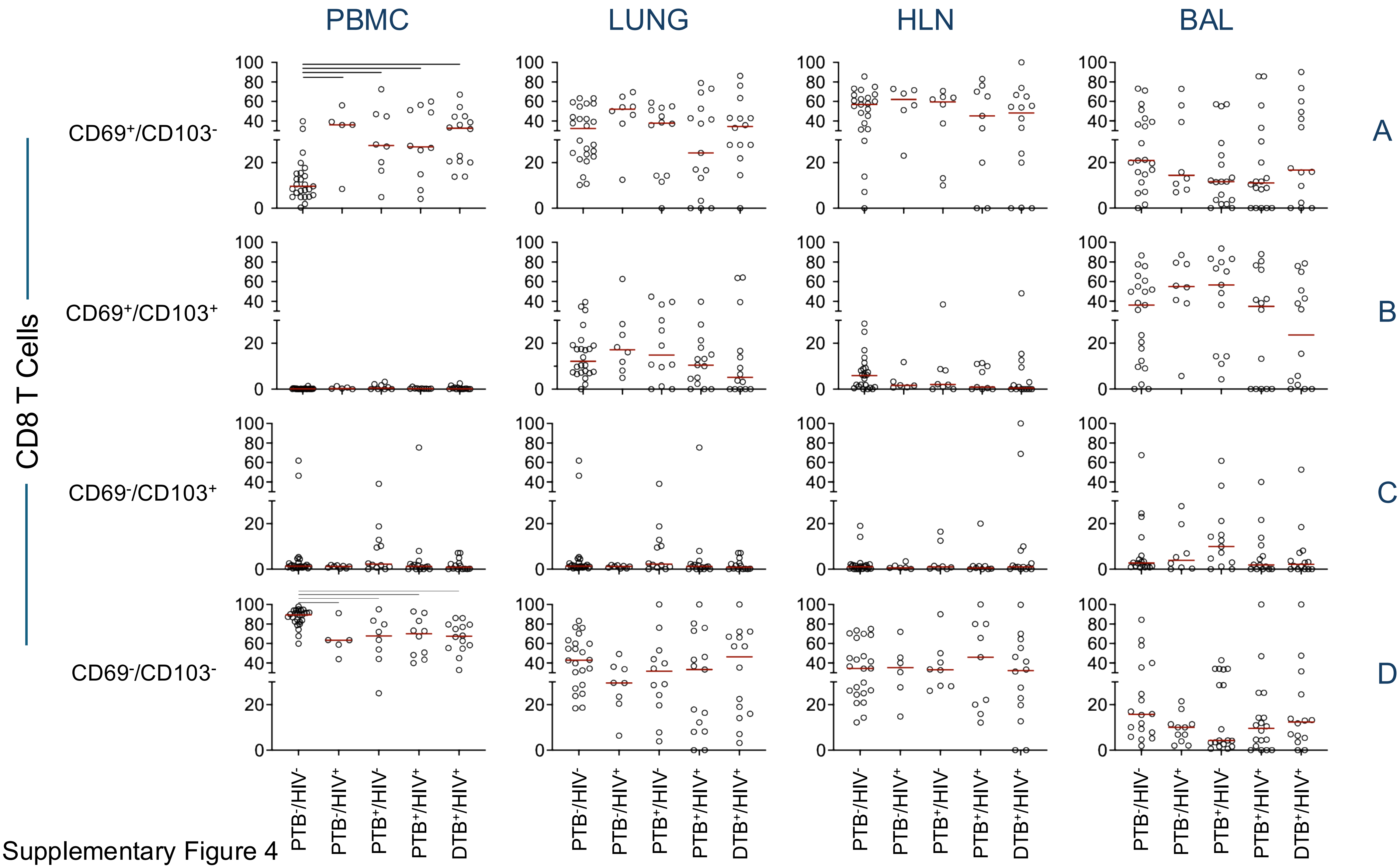


Supplementary Figure 4
